# Assessment of the evidence yield for the calibrated PP3/BP4 computational recommendations

**DOI:** 10.1101/2024.03.05.24303807

**Authors:** Sarah L. Stenton, Vikas Pejaver, Timothy Bergquist, Leslie G. Biesecker, Alicia B. Byrne, Emily Nadeau, Marc S. Greenblatt, Steven Harrison, Sean Tavtigian, Predrag Radivojac, Steven E. Brenner, Anne O’Donnell-Luria, ClinGen Sequence Variant Interpretation Working Group

**Affiliations:** Program in Medical and Population Genetics, Broad Institute of MIT and Harvard, Cambridge, MA 02142, USA; Division of Genetics and Genomics, Boston Children's Hospital, Harvard Medical School, Boston, MA 02115, USA; Institute for Genomic Health, Icahn School of Medicine at Mount Sinai, New York, NY 10029, USA; Department of Genetics and Genomic Sciences, Icahn School of Medicine at Mount Sinai, New York, NY 10029, USA; Center for Precision Health Research, National Human Genome Research Institute, National Institutes of Health, Bethesda, MD 20892, USA; Department of Medicine and University of Vermont Cancer Center, University of Vermont, Larner College of Medicine, Burlington, VT 05405, USA; Ambry Genetics, Aliso Viejo, CA, USA; Department of Oncological Sciences, Huntsman Cancer Institute, University of Utah School of Medicine, Salt Lake City, UT 84112, USA; Khoury College of Computer Sciences, Northeastern University, Boston, MA 02115, USA; Department of Plant and Microbial Biology and Center for Computational Biology, University of California, Berkeley, Berkeley, CA 94720, USA

## Abstract

**Purpose:** To investigate the number of rare missense variants observed in human genome sequences by ACMG/AMP PP3/BP4 evidence strength, following the calibrated PP3/BP4 computational recommendations.

**Methods:** Missense variants from the genome sequences of 300 probands from the Rare Genomes Project with suspected rare disease were analyzed using computational prediction tools able to reach PP3_Strong and BP4_Moderate evidence strengths (BayesDel, MutPred2, REVEL, and VEST4). The numbers of variants at each evidence strength were analyzed across disease-associated genes and genome-wide.

**Results:** From a median of 75.5 rare (≤1% allele frequency) missense variants in disease-associated genes per proband, a median of one reached PP3_Strong, 3-5 PP3_Moderate, and 3-5 PP3_Supporting. Most were allocated BP4 evidence (median 41-49 per proband) or were indeterminate (median 17.5-19 per proband). Extending the analysis to all protein-coding genes genome-wide, the number of PP3_Strong variants increased approximately 2.6-fold compared to disease-associated genes, with a median per proband of 1-3 PP3_Strong, 8-16 PP3_Moderate, and 10-17 PP3_Supporting.

**Conclusion:** A small number of variants per proband reached PP3_Strong and PP3_Moderate in 3,424 disease-associated genes, and though not the intended use of the recommendations, also genome-wide. Use of PP3/BP4 evidence as recommended from calibrated computational prediction tools in the clinical diagnostic laboratory is unlikely to inappropriately contribute to the classification of an excessive number of variants as Pathogenic or Likely Pathogenic by ACMG/AMP rules.

## INTRODUCTION

Genetic testing identifies many variants of uncertain significance (VUS), of which the majority of coding variants are missense (non-synonymous).^1^ Limited availability of functional data means that it is often necessary to turn to computational *in silico* prediction tools for evidence of deleteriousness. The American College of Medical Genetics and Genomics and the Association for Molecular Pathology (ACMG/AMP) provide a sequence variant classification (SVC) framework to combine distinct lines of evidence of pathogenicity or benignity of varying strengths to reach a final variant classification (Benign [B], Likely Benign [LB], VUS, Likely Pathogenic [LP], or Pathogenic [P]). In the 2015 recommendations, *in silico* evidence (PP3 and BP4) was capped at “Supporting” for or against pathogenicity.^2^ Furthermore, no explicit recommendations concerning the prediction tools or thresholds to be used were specified, enabling non-standardized application of criteria and resulting in inconsistencies in variant classification between clinical diagnostic laboratories.^3^

Recently, Pejaver et al., (2022) refined the use of computational prediction tools to provide evidence of pathogenicity using the Bayesian adaptation of the ACMG/AMP framework.^4,5^ For 13 computational prediction tools frequently used in clinical workflows, evidence-based calibrated thresholds were introduced corresponding to “Supporting,” “Moderate,” “Strong,” and “Very Strong” PP3/BP4 evidence strengths, and also defined an indeterminate range. These thresholds demonstrated that the initial framework underweighted evidence from computational prediction tools, as many had the ability to provide evidence beyond “Supporting” strength.

Since the release of the PP3/BP4 recommendations, we have received questions from users regarding the key steps to implementation, calling for practical guidance on the intended use of the PP3/BP4 recommendations for variant curation in disease-associated genes (see **Box 1**). In particular, concerns have arisen due to the impression that an excessive number of variants are reaching PP3_Strong. Here, by demonstrating the level of PP3/BP4 evidence allocated to rare missense variants in the genome sequences of patients with rare disease, we specifically aimed to address these concerns.

### Box 1.

Key steps in implementing the PP3/BP4 missense variant recommendations

#### How should PP3/BP4 evidence be used for missense variants?

- Only apply PP3/BP4 evidence in genes where missense variants are known to cause disease
- Use a single computational prediction tool, preferably one able to reach PP3_Strong and BP4_Moderate
- Select the prediction tool in advance of seeing the scores, and preferably before knowing the results of any other line of evidence (i.e. do not “cherry pick” a prediction tool)
- Use established thresholds unless or until there is superseding gene or region-specific guidance

#### How can computational PP3 evidence be combined with other ACMG/AMP codes?

- Use the PP3/BP4 evidence within the ACMG/AMP rules for variant classification, and implement all updated recommendations together
- Code combination must avoid double-counting of evidence, for example:
  - The combined evidence strength of PP3 and PM1 is capped at “Strong”
  - Prediction tools that explicitly incorporate allele frequency should not be combined with independent allele frequency evidence

#### How can the calibrations be customized by expert panels to specific genes or regions?

- Apply the prediction tool that performs best for the gene(s) or region
- If there is evidence that the prediction tool yields inappropriate predictions for a specific gene or region, well-informed judgment may be used to adjust PP3/BP4 evidence
- For genes with sufficient number of benign and pathogenic missense variants, it may be possible to perform gene (or region) specific calibration

#### Can the calibration of these methods be trusted?

- PP3/BP4 are empirically calibrated evidence codes
- Confounders that could be addressed directly were eliminated
- As with any approach, it is expected that the evidence strength provided will be too high or too low for some variants when applying the calibrated PP3/BP4 codes
- The calibrated codes have been extensively validated, including in this study

#### What are some of the limitations to the calibration?

- Variants used for the calibration may not be representative of novel variants to be classified
- Computational prediction tools were assumed not to have had a major role in the classification of variants used in the calibration
- The calibration provides the evidence strength, on average, across the thousands of genes assessed; however, it is a probability that will vary across genes

#### Will more calibrations need to be performed in the future?

- New and revised methods will require independent calibration

More detailed answers to these questions are provided in the **Supplemental Material**

## METHODS

### Study participants and data

Genome sequencing (GS) data were obtained from the Rare Genomes Project (RGP) at the Broad Institute of MIT and Harvard.^6^ All participants signed informed consent including the use of data for research purposes (Mass General Brigham IRB protocol 2016P001422). Participant demographics are displayed in **Table S1**. Sequencing was performed on DNA purified from blood by the Broad Institute Genomics Platform on an Illumina sequencer to 30x average depth. Raw sequence reads were aligned to the GRCh38 reference genome. Variants were called with GATK version 4.1.8.0^7^ in the form of single nucleotide variants (SNVs) and small insertions/deletions (indels). Variants were filtered at the site-level with GATK Variant Quality Score Recalibration (VQSR).

### Missense variant extraction and annotation

Missense variants identified by the Ensembl Variant Effect Predictor (VEP)^8^ using MANE Select transcripts^9^ were extracted from the GS data. Only variants with genotype quality ≥40, depth ≥10, and allele balance ≥0.2 were retained for analyses. Allele frequency (AF) thresholds of ≤5% and ≤1% global and population-max “popmax” AF in gnomAD v3.1.2 genomes were applied (the highest allele frequency for non-bottlenecked populations).^10^ Precomputed scores from four *in silico* (meta)predictors that were able to reach PP3_Strong and BP4_Moderate in the Pejaver et al. calibration were included in the analysis. BayesDel (without minor allele frequency),^11^ REVEL,^12^ and VEST4^13^ were annotated using the dbNSFP4.4a database.^14^ MutPred2 scores were also generated.^15^ For transcript-specific predictors, MutPred2, REVEL, and VEST4, the MANE Select transcript was used for score annotation.

Using recommended thresholds,^4^ PP3/BP4 evidence strength per variant was annotated for each prediction tool. The number of missense variants summed across probands and per proband by prediction tool and evidence strength was assessed in disease-associated genes, classified as “Definitive,” “Strong,” or “Moderate” in the Gene Curation Coalition (GenCC) Database (last accessed Jul 21, 2023) (3,424 genes - 1,004 autosomal dominant only [AD-only], 1,903 autosomal recessive only [AR-only], 517 other [includes gene that are both AD and AR])^16^ and genome-wide. These methods were also repeated for missense variants according to the VEP “most severe consequence” across all transcripts (see **Supplemental Material**).

### Statistical analyses

Proportions between two groups were compared with two-tailed binomial tests with Bonferroni correction for multiple testing. Bootstrap resampling with replacement (1,000 iterations) was performed to provide a 95% confidence interval (CI) for the mean.

## RESULTS

### Detection of missense variants in disease-associated genes

The GS dataset included 300 probands with rare disease. Across protein-coding genes genome-wide, a median of 8,781.5 variants per proband (range 8,383-10,616) passing QC thresholds were detected in MANE Select transcripts. Applying a ≤1% AF threshold in the gnomAD v3 genomes dataset, we found 75,384 unique missense variants across 15,566 genes (median 321 per proband, range 244-847). Within GenCC Moderate, Strong, and Definitive disease-associated genes, the number of unique variants dropped to 17,789 across 2,899 genes, and a median of 75.5 variants per proband (range 53-186). Variant counts following each step in QC and AF filtering are displayed in **Table S2**.

### PP3/BP4 evidence strength of missense variants in disease-associated genes

A median of one variant (mean 0.8-1) per proband reached PP3_Strong per analyzed prediction tool, 3-5 variants (mean 3.4-4.9) reached PP3_Moderate, and 3-5 (mean 3.6-5.2) reached PP3_Supporting (**Table 1, Figure 1A**). Summed across all probands, 227-313 PP3_Strong variants were found in a total of 153-196 disease-associated genes, accounting for 0.96-1.3% of all analyzed variants (BayesDel 1.3% [95% CI 1.321-1.330], MutPred2 1.0% [95% CI 1.037-1.044], REVEL 0.96% [95% CI 0.957-0.966], VEST4 1.1% [95% CI 1.113-1.114]). PP3_Moderate-Strong variants were more frequent in autosomal recessive (AR) genes than in autosomal dominant (AD) genes (p-value ≤0.0001 for all prediction tools, two-tailed binomial test with Bonferroni correction) **(Figure 1B** and **Figure S1**). ClinVar provides classifications for 54-72% of the unique variants with PP3_Strong evidence per prediction tool, of which 12-29% are currently reported as P/LP, 63-79% as VUS or Conflicting interpretations of pathogenicity, and 7-10% as B/LB (**Figure S2**). The majority of analyzed variants were allocated BP4 evidence (median 41-49 per proband, 53-64% of all analyzed variants) or were indeterminate (median 17.5-19 per proband, 23-25% of all analyzed variants) (**Figure S3**). Using a preliminary calibration, the newly released prediction tool AlphaMissense^17^ was generally consistent with each of these figures (data not shown).

**Table 1.** Number of rare (≤1% AF) missense variants in disease-associated genes per proband by ACMG PP3/BP4 evidence strength within MANE Select transcripts.

| Table 1. Number of rare ($\leq 1\%$ AF) missense variants in disease-associated genes per proband by ACMG PP3/BP4 evidence strength within MANE Select transcripts | | | | | | | | |
| --- | --- | --- | --- | --- | --- | --- | --- | --- |
| ACMG/AMP evidence code | Computational prediction tool |  |  |  |  |  |  |  |
|  | BayesDel |  | MutPred2 |  | REVEL |  | VEST4 |  |
|  | Median (range) | Mean (sd) | Median (range) | Mean (sd) | Median (range) | Mean (sd) | Median (range) | Mean (sd) |
| PP3_Strong | 1 (0-4) | 1 (1) | 1 (0-4) | 0.8 (0.9) | 1 (0-4) | 0.8 (0.9) | 1 (0-4) | 0.9 (0.9) |
| PP3_Moderate | 4 (0-10) | 4.1 (1.9) | 4 (0-12) | 4 (2) | 3 (0-9) | 3.4 (1.9) | 5 (0-11) | 4.9 (2) |
| PP3_Supporting | 4 (0-13) | 4.6 (2.2) | 4 (0-11) | 3.8 (2) | 3 (0-9) | 3.6 (1.8) | 5 (0-13) | 5.2 (2.3) |
| Indeterminate | 18 (7-35) | 18.6 (4.8) | 19 (8-44) | 19.7 (5.2) | 18 (8-36) | 18.4 (5.1) | 17.5 (4-31) | 17.9 (4.4) |
| BP4_Supporting | 17 (8-48) | 17.8 (5.1) | 18 (9-39) | 18.2 (5.1) | 11 (3-23) | 11 (3.6) | 11 (2-40) | 11.3 (4.2) |
| BP4_Moderate | 31 (15-91) | 32.3 (9.8) | 31 (14-96) | 32.2 (9.7) | 28 (14-84) | 29.3 (8.7) | 32 (15-91) | 32.8 (10.1) |
| BP4_Strong | - | - | 0 (0-2) | 0.1 (0.3) | 1 (0-11) | 1.6 (1.6) | - | - |
| BP4_Very Strong | - | - | - | - | 0 (0-1) | 0 (0.2) | - | - |
| No score | 0 (0-3) | 0.3 (0.6) | 0 (0-0) | 0 (0) | 10 (2-28) | 10.7 (3.6) | 5 (1-19) | 5.8 (2.8) |

“-” indicates that the given prediction tool is not able to provide BP4 evidence of this strength.

**Figure 1.**
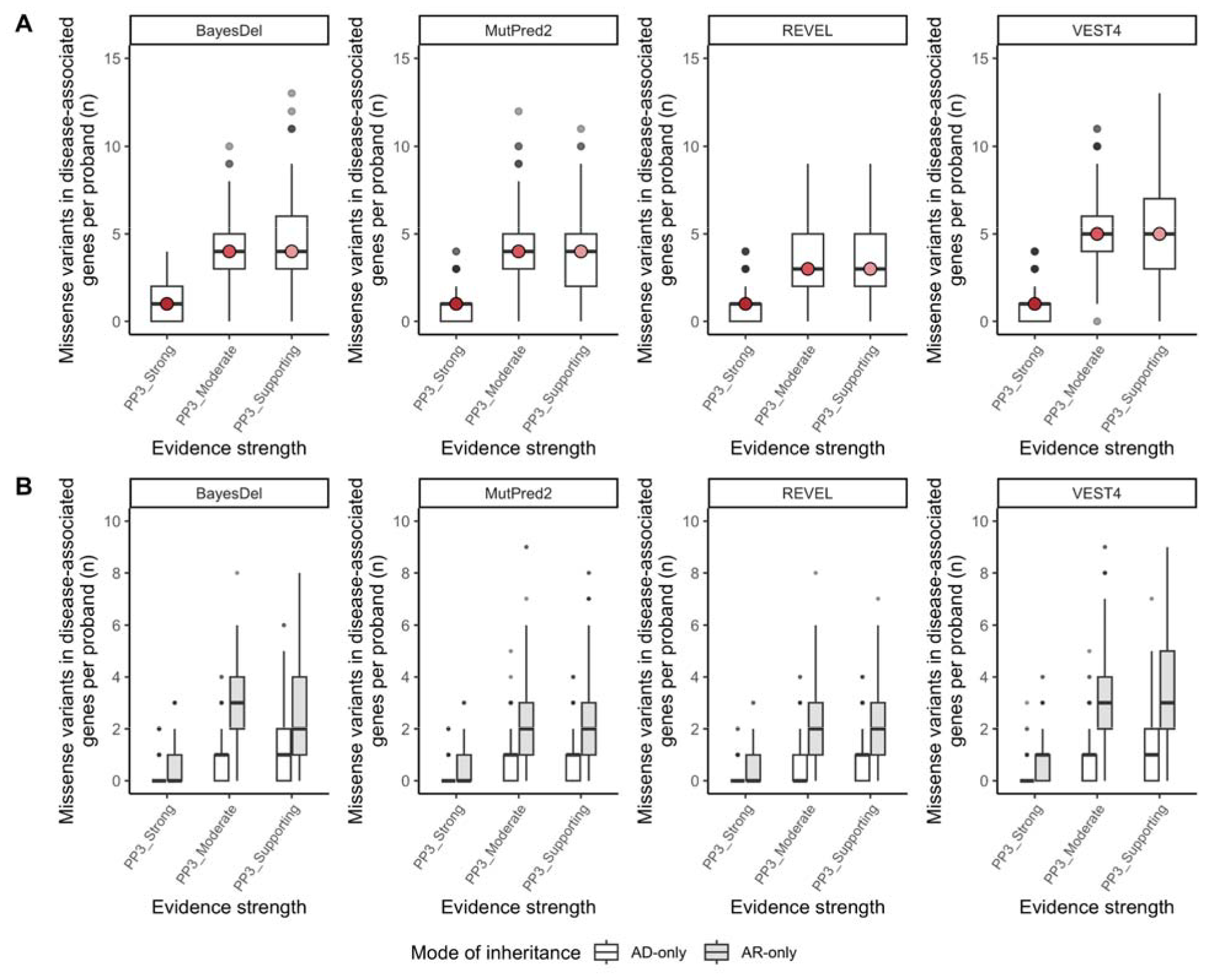
**A**. Rare (≤1% AF) missense variants in disease-associated genes per proband by PP3 evidence strength for analyzed computational prediction tools. **B**. Rare (≤1% AF) missense variants in disease-associated genes with PP3 evidence per proband by evidence strength and reported mode of inheritance (AD-only and AR-only) for analyzed computational tools. Boxplots correspond to the first, second, and third quartile of data, with whiskers denoting 1.5 × IQR. Outliers are displayed as individual points.

Using a less stringent AF threshold of ≤5% resulted in a subtle increase in variants with PP3 evidence in disease-associated genes (median = 1 PP3_Strong, median = 4-6 PP3_Moderate, median = 5-6 PP3_Supporting) (**Table S3**). Using the VEP “most severe consequence” across all transcripts to detect variants, a rare disease analysis approach that is sometimes used to increase the detection of potentially deleterious missense variants in alternative transcripts versus using only MANE Select transcripts, we also did not see many more variants reaching PP3_Supporting-Strong in disease-associated genes (median = 1 PP3_Strong, median = 3-5 PP3_Moderate, median = 4-5 PP3_Supporting) (**Table S4**).

Although the ACMG/AMP SVC framework is intended to be used in genes associated with Mendelian disease, we also analyzed variants genome-wide and found 1-3 PP3_Strong, 8-16 PP3_Moderate, and 10-17 PP3_Supporting variants per proband (**Table S5**). Summed across all probands, a total of 447-847 variants with PP3_Strong evidence were found in 317-587 protein-coding genes genome-wide, accounting for 0.442-0.837% of all analyzed variants (BayesDel 0.611% [95% CI 0.610-0.613%], MutPred2 0.837% [95% CI 0.835-0.838%], REVEL 0.442% [95% CI 0.441-0.444%], VEST4 0.792% [95% CI 0.791-0.794%]). This equates to a 2-3.5-fold (mean 2.6-fold) higher number than found at the same AF threshold in disease-associated genes only.

## DISCUSSION

The use of computational prediction tools to provide evidence of pathogenicity and benignity within the ACMG/AMP framework was recently refined by Pejaver et al.^4^ and certain prediction tools were found capable of reaching “Strong” and “Very Strong” evidence for PP3 and BP4 codes, respectively. These changes were expected to have important implications for the final classification of missense variants in the clinical diagnostic setting, given that previously the codes were capped at “Supporting” and could only be applied if “Multiple” lines of computational evidence support a deleterious effect on the gene or gene product.^2^

Through various scientific meetings and interactions following release of the recommendations, concerns were raised due to the impression that an excessive number of PP3_Strong variants are generated. To explore these concerns, we assessed the observed number of rare missense variants by PP3/BP4 evidence strength in the genome sequences of 300 research participants with rare disease.

In our analyses, at ≤1% AF, a standard threshold in rare disease analysis, we found a median of one PP3_Strong variant per individual (range 0-4) across ∼950 of over 3,400 analyzed disease-associated genes. Most variants had evidence of benignity or no evidence (indeterminate). The rate of PP3_Strong variants amongst all rare missense variants in disease-associated genes per individual ranged from 0.96-1.3%, which is consistent with previous reports for variants similarly sampled from gnomAD (1.4-1.7%).^4^ The PP3_Moderate-Strong variants were more frequently found in genes associated with disorders having AR inheritance, in which heterozygous deleterious variants can be expected that are non-diagnostic in the individual, compared to genes associated with disorders having AD inheritance, where non-diagnostic variants may represent false positives. Moreover, for PP3_Strong variants with ClinVar classifications available, ≤10% were classified as B/LB. These figures suggest that there is a low risk of attributing PP3_Strong evidence to a high number of false-positive variants per individual, and also confirm that gnomAD is an appropriate reference set for score calibration for application in variant classification.

To better understand why users reported an excess of PP3_Strong variants, we also extended our analyses to more frequent variants up to 5% AF, the threshold for stand-alone evidence of benignity in the ACMG/AMP guidance, and to variants that are missense on alternative transcripts (VEP “most severe consequence”). These analyses did not result in a considerable increase in the number of PP3_Strong variants. Furthermore, though Pejaver et al. made no recommendation about running computational prediction tools genome-wide, given that the thresholds are calibrated for disease-associated genes only, we applied the same thresholds to variants genome-wide. We found an approximately 2.6-fold increase in the number of PP3_Strong variants genome-wide compared to within disease-associated genes only, consistent with the genome having ∼5-fold as many genes as covered by ACMG/AMP classification rules and the prior for pathogenicity genome-wide being ∼5-fold lower (∼1%)^15,18^ than for disease-associated genes (∼4.5%)^4^.

Importantly, deleterious *in silico* prediction does not equate to pathogenicity and, in the absence of additional evidence, one line of “Strong” evidence from the PP3 code classifies a variant as a VUS in the ACMG/AMP framework. In the case that a variant does reach P or LP classification in combination with other codes, there is a 99% or 90% posterior probability of pathogenicity, respectively, which implies that 1-10% of variants may not actually be causative of disease. The PP3/BP4 codes should be used within the framework of the ACMG/AMP recommendations including the updates that have been made by ClinGen to determine the pathogenicity of a variant. Code combination does, however, require great care and there are a number of important caveats. In particular, (meta)predictors may use data partially captured by other codes, notably key domains and critical residues and population AF, increasing the risk of double-counting of evidence (see **Supplemental Material** for further recommendations on code combination).

The PP3/BP4 calibration by Pejaver et al. does have limitations. It was performed on variants classified in the past several years that were not used in the training sets of the analyzed prediction tools and may be non-representative of novel variants to be classified. Moreover, computational prediction tools were assumed not to have played a major role in the classification of the variants used for the calibration. Given these limitations, we appeal to

diagnostic labs to report the computational prediction tool and version used for PP3/BP4 evidence, both to determine the impact the PP3/BP4 recommendations are having on the final classification of missense variants and to ensure that we are able to continue to evaluate the performance of computational predictors in the future.

## CONCLUSION

A small number of variants per rare disease proband reached PP3_Strong and PP3_Moderate in disease-associated genes following the calibrated PP3/BP4 computational recommendations. Computational methods are therefore unlikely to inappropriately classify variants as P/LP by ACMG/AMP rules in the clinical setting by the ACMG/AMP framework.

## Data Availability

All data produced in the present work are contained in the manuscript.

## ETHICS DECLARATION

All participants signed informed consent including the use of data for research purposes (Mass General Brigham IRB protocol 2016P001422).

## ClinGen Sequence Variant Interpretation Working Group

Leslie G. Biesecker (co-chair), Steven M. Harrison (co-chair), Ahmad A. Tayoun, Jonathan S. Berg, Steven E. Brenner, Garry R. Cutting, Sian Ellard, Marc S. Greenblatt, Peter Kang, Izabela Karbassi, Rachel Karchin, Jessica Mester, Anne O’Donnell-Luria, Tina Pesaran, Sharon E. Plon, Heidi L. Rehm, Natasha T. Strande, Sean V. Tavtigian, and Scott Topper.

