## Supplemental for "Assessment of the evidence yield for the calibrated PP3/BP4 computational recommendations"

**SUPPLEMENTAL MATERIAL**

**Key steps in implementing the PP3/BP4 missense variant recommendations**

**Q1. How should PP3/BP4 evidence be used?**

Pejaver et al., calibrated 13 computational tools to provide evidence of pathogenicity and benignity within the ACMG/AMP framework.^1^ The recommended thresholds thereby only apply to genes where missense variants are known to cause disease and are *only* suitable for use in the process of variant classification within known disease genes in the context of the ACMG/AMP rules. The recommendation does not apply to entire exomes or genomes. For the allocation of PP3/BP4 evidence, a single computational tool should be used, preferably one able to reach PP3_Strong and BP4_Moderate (BayesDel, MutPred2, REVEL, or VEST4) and this tool should be selected in advance of seeing the score to avoid cherry picking the “best” scores and ideally, before seeing any other evidence. The established calibrated thresholds should be used unless or until there is superseding gene/region-specific guidance.

**Q2. How can computational PP3 evidence be combined with other ACMG/AMP codes?**

PP3/BP4 evidence should be used within the ACMG/AMP rules for variant classification, and should be implemented together with all updated recommendations (e.g., no longer using the PP5/BP6 codes^2^, along with altered use of the PM1 and PM2 codes, described below). The ACMG/AMP codes must be combined carefully, as there are a number of important caveats to consider to avoid double-counting.

The first category of codes to combine carefully are those based on key domains and critical residues (PM1, PS1, and PM5). The updated recommendations^1^ support combining PP3 with PM1 (located in a mutational hot spot and/or critical and well-established functional domain [e.g., active site of an enzyme] without benign variation) to an evidence strength capped at “Strong”. This may, however, not be justifiable, and indeed no calibration of this combination of evidence was performed to date. PS1 (same amino acid change as a previously established pathogenic variant regardless of nucleotide change) and PM5 (novel missense change at an amino acid residue where a different missense change determined to be pathogenic has been seen before) partially overlap with PP3 conceptually, with the important distinction that both require evidence of an abnormal human phenotype.

The second category of codes are those based on population AF (PM2, BS1, and PS4). PM2 (absent from controls [or extremely low AF if AR]) and BS1 (AF is greater than expected for the disorder) should only be used in combination with predictions from tools that do not explicitly incorporate low population AF when weighing variant pathogenicity and should use the current recommendations to downgrade PM2 to “Supporting”.^3^ Indeed, there is evidence that current databases are not of sufficient size to reach saturation that would support the use of the PM2 code at all.^4^ As in the case of PS1 and PM5, the PS4 code (prevalence of the variant in affected individuals is significantly increased compared with the prevalence in controls) demonstrates an effect on human phenotype and undoubtedly buttresses a pure *in silico* prediction. Overall, given that these ACMG/AMP codes are, to some extent, confounded, more sophisticated guidance on their combination is required.

Foreseeably, migration to a point-based variant classification system could enable the use of fractional points for PP3/BP4 on a continuous scale that corresponds to the continuous output of the tool. However, we caution against such an approach, as the current limitations of the calibration mean the precision is not sufficient to warrant confident assignment of fractional points and further work is needed to properly assess these possibilities further.

**Q3. How can the calibrations be customized to specific genes/regions?**

The indicated scores provide the evidence strength, on average, across the thousands of genes assessed. However, there is variability. Some regions/domains, genes, or gene classes would be expected to systematically have too many variants called at a given PP3/BP4 strength, balancing those that have too few.

Ideally, gene/region-specific thresholds would be defined. However, due to the requirement to calibrate tools with data that was not used in the methods’ developments, the reduced scale of the analyses makes using the methods implemented by Pejaver et al. essentially impossible without overfitting. This remains an area for further method development. Therefore, unless or until there is gene/region-specific superseding guidance, such as the release of refined thresholds from a ClinGen Variant Curation Expert Panel (VCEP),^5^ we recommend the use of a single tool, at the recommended thresholds, able to reach PP3_Strong and BP4_Moderate genome-wide. As stated above, the tool should be selected in advance and ideally, before seeing any other evidence. If the selected tool is found to make an excessive number of predictions at too high a degree of evidence in a specific gene/region, VCEPs may use informed judgment to adjust PP3/BP4 evidence. We do not recommend altering the evidence strength of the tools unless there is exceptional expert judgment that warrants this.

**Q4. Can the calibration of these methods be trusted?**

We think so, on average. The PP3/BP4 calibration was rigorously performed to eliminate confounders that could be addressed directly (e.g., removing variants that were part of methods’ training sets), making it one of the most empirically calibrated ACMG/AMP codes.

**Q5. What are some of the limitations to the calibration?**

The greatest concern about the reliability of the PP3/BP4 thresholds defined by Pejaver et al. is that the calibration was performed only on variants identified in the past several years and that were not used in tool training sets. To what extent newly identified variants requiring classification differ from these is unknown, and may impact calibration accuracy. For example, variants that have been classified to date may be easier to identify than others and non-representative of missense variants as a whole. We do not have a good way to identify in what way such variants are non-representative and it is not clear whether the calibration produces too many false positives or negatives. Moreover, tools were calibrated for the canonical transcript only, and not for alternative transcripts that are often considered in clinical variant interpretation. We also assumed that computational methods did not have a major role in the classification of the evaluation set, which was warranted as PP3/BP4 had previously only provided evidence up to “Supporting”, and thus provided minor evidence relative to other codes.

**Q6. Will more calibrations need to be performed in the future?**

While we do not expect recalibration will be needed for the existing methods, new methods are emerging which show promise for more precise and effective predictions of variant pathogenicity, according to the CAGI experiment^6^ and publications.^7^ Evaluations that take steps to avoid circularity should be performed to confirm this.

**SUPPLEMENTAL METHODS**

**Missense variant extraction and annotation using “most severe consequence” Variant Effect Prediction.** Missense variants were extracted from the GS data using the “most severe consequence” across all transcripts according to the Ensembl Variant Effect Predictor (VEP), an approach that is used in some analysis pipelines that will increase the number of damaging missense variants over just using MANE select or Ensembl canonical transcripts.^8^ Only high quality variants defined as those with genotype quality ≥40, depth ≥10, and allele balance ≥0.2 were retained for analysis. An allele frequency (AF) threshold of ≤1% global and population-max “popmax” AF in gnomAD v3.1.2 genomes was applied (the highest allele frequency for non-bottlenecked populations).^9^ Precomputed scores from the *in silico* (meta)predictors that reached PP3_Strong and BP4_Moderate in the calibration were included here. BayesDel (without minor allele frequency),^10^ REVEL,^11^ and VEST4,^12^ were annotated using the database of human nonsynonymous SNPs and their functional predictions (dbNSFP4.4a).^13^ MutPred2 scores were generated for the variants considered in this study.^14^ For the transcript-specific predictors, MutPred2, REVEL, and VEST4, the MANE Select transcript was used for score annotation, when applicable. When the transcript with the “most severe consequence” was not the MANE Select transcript, all applicable alternative transcripts were annotated and the highest score across transcripts was retained. Using thresholds recommended by Pejaver et al,^1^ the PP3/BP4 evidence strength according to each of the tools was annotated per variant. The number of missense variants per proband by tool and evidence strength was counted in disease associated genes, classified as “Definitive”, “Strong”, or “Moderate” in the Gene Curation Coalition (GenCC) Database (last accessed Jul 21, 2023) (3,424 genes)^15^ and genome-wide.

**SUPPLEMENTAL FIGURES**

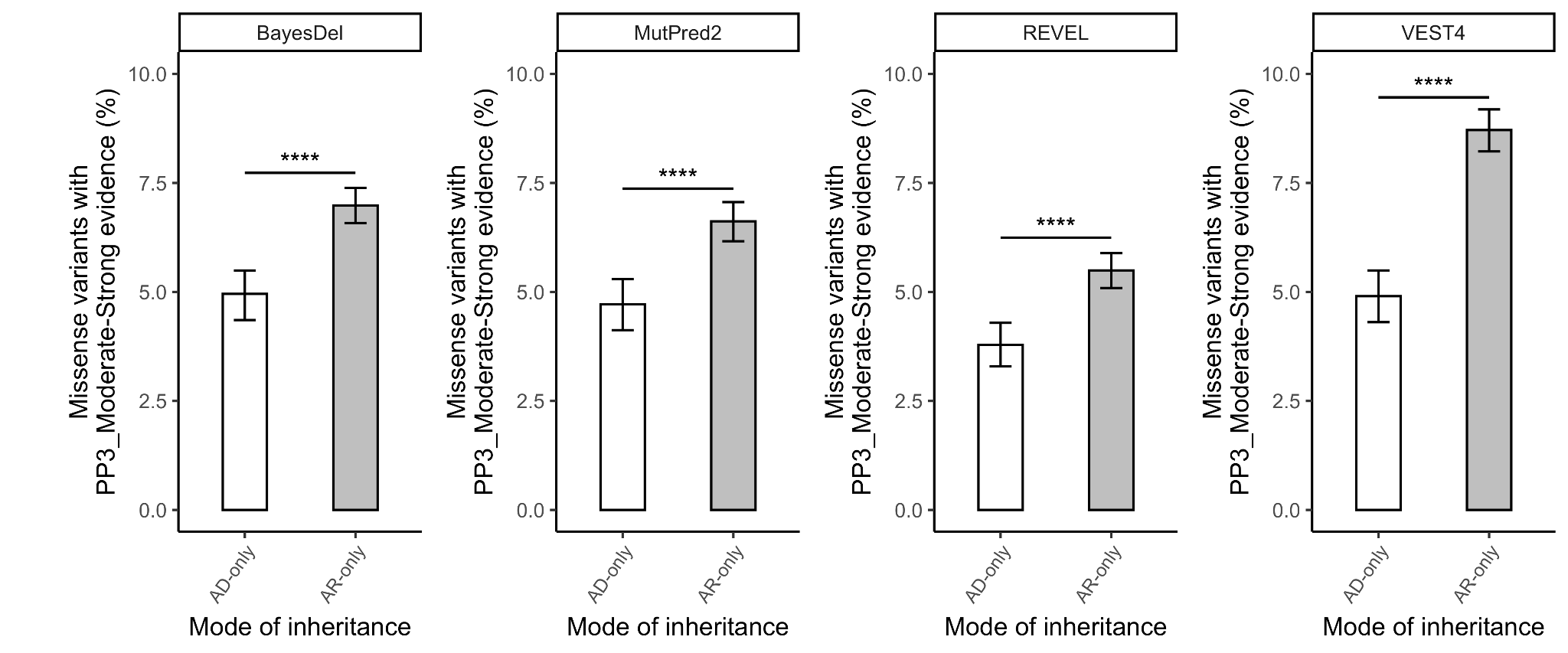

**Figure S1.** Percentage of analyzed variants in disease-associated genes with either AD-only or AR-only reported mode of inheritance across all probands with PP3_Moderate-Strong evidence. Groups were compared with a two-tailed binomial test, p-values Bonferroni corrected for multiple testing (ns, >0.05; *, ≤0.05; **, ≤0.01; ***, ≤0.001; ****, ≤0.0001). Data displayed with 95% confidence intervals.

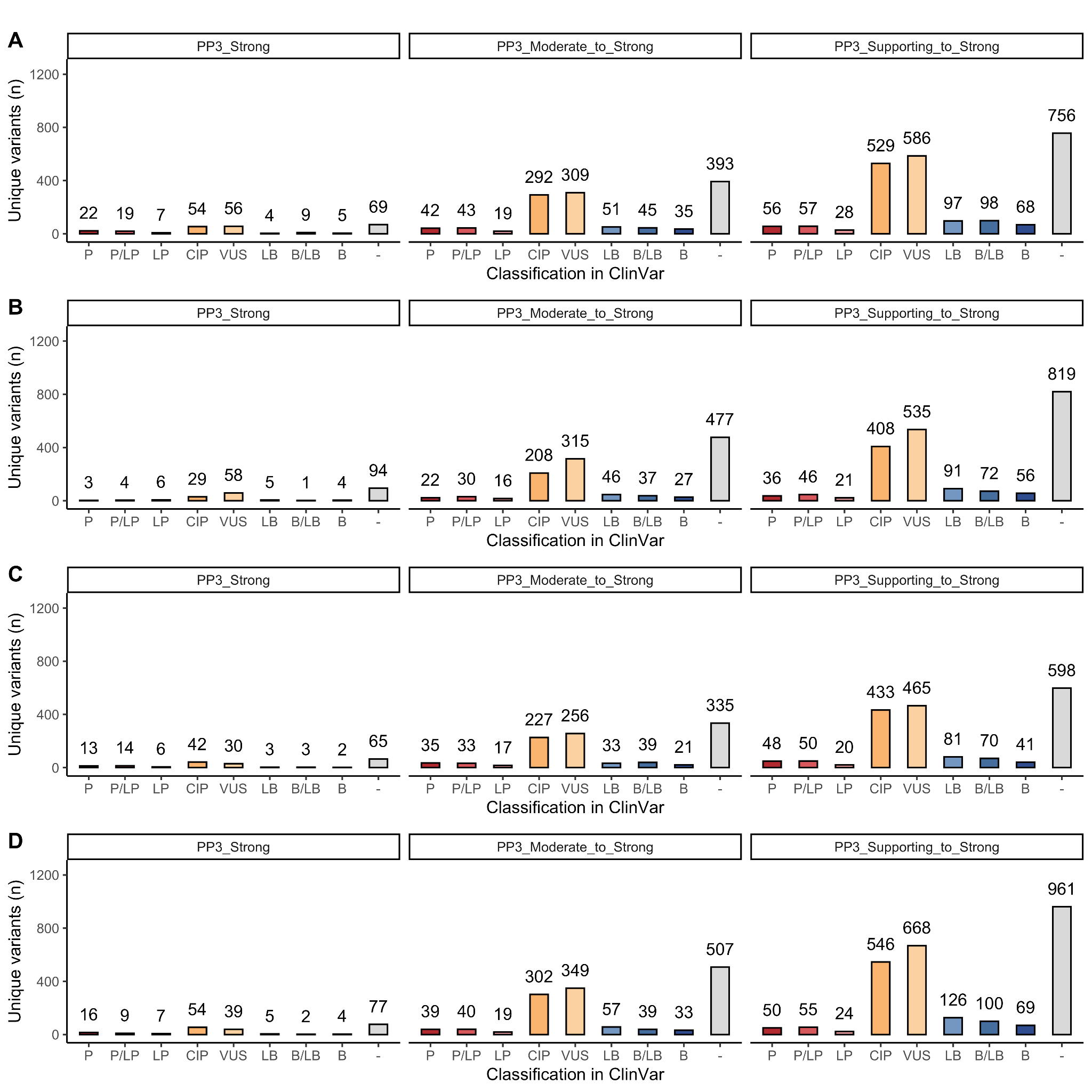

**Figure S2. Classification in ClinVar of unique variants reaching PP3 evidence strength by each analyzed tool. A.** BayesDel. **B.** MutPred2. **C.** REVEL. **D.** VEST4. Middle and right panels are cumulative counts. P, pathogenic; LP, likely pathogenic; CIP, conflicting interpretations of pathogenicity; VUS, variant of uncertain significance; LB, likely benign; B, benign. “-” indicates that the variant does not have a classification in ClinVar (last accessed, Aug 28th 2023).

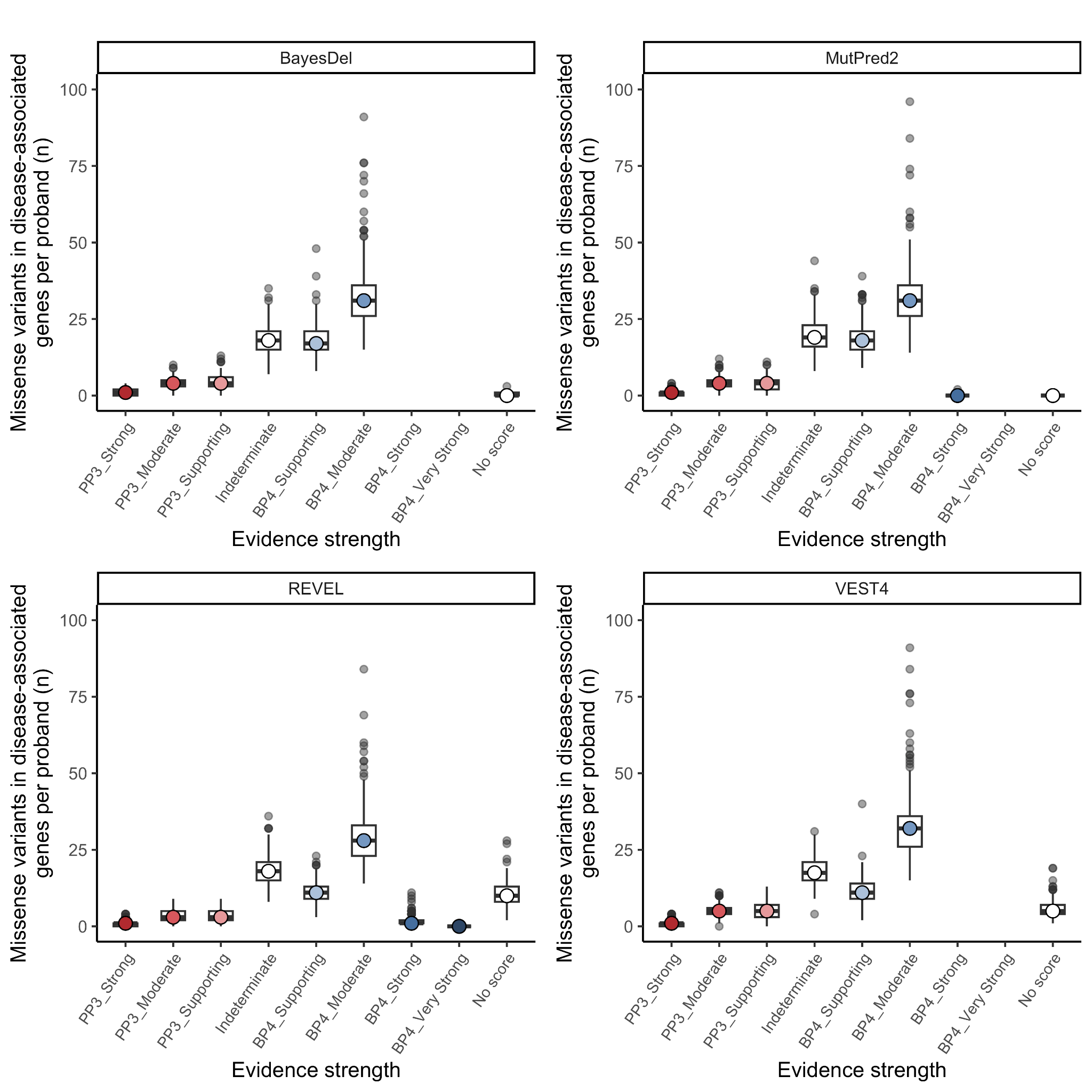

**Figure S3.** Rare (≤1% AF) missense variants in disease-associated genes per proband by PP3/BP4 evidence strength for analyzed computational tools. Boxplot corresponding to the first, second, and third quartile of data, with whiskers denoting 1.5 × IQR. Outliers are displayed as individual points.

**SUPPLEMENTAL TABLES**

| **Table S1. Participant demographics** | | |
| --- | --- | --- |
| **Sex** | Female | 142 (47%) |
|  | Male | 158 (53%) |
| **Ancestry** | Ashkenazi Jewish | 21 (7%) |
|  | Black or African American | 17 (6%) |
|  | East Asian | 5 (2%) |
|  | Hispanic or Latino | 23 (8%) |
|  | Other | 50 (17%) |
|  | South Asian | 5 (2%) |
|  | White | 179 (60%) |

| **Table S2. Summary of missense variant counts by filtering step** | | | | | |
| --- | --- | --- | --- | --- | --- |
| **Filtering step** | **Total variants (n)** | **Unique variants (n)** | **Genes with variants (n)** | **Variants per proband (n)** | |
|  |  |  |  | Median (range) | Mean (sd) |
| **All** | 3,338,480 | 281,104 | 17,872 | 10,996 (10,490-13,191) | 11,128 (480) |
| **GQ ≥40, DP ≥10, plus AB ≥0.2** | 2,670,086 | 257,508 | 17,564 | 8,781.5 (8,383-10,616) | 8,900 (421) |
| **≤5% AF** | 207,208 | 97,836 | 16,113 | 640.5 (544-1,540) | 691 (169) |
| **≤1% AF** | 101,319 | 75,384 | 15,566 | 321  (244-847) | 338 (65) |
| **Disease-**  **associated gene** | 23,629 | 17,789 | 2,899 | 75.5  (53-186) | 79 (16) |

GQ, genotype quality; DP, depth; AB, allele balance; AF, allele frequency.

| **Table S3. Number of rare (≤5% AF) missense variants in disease-associated genes per proband by ACMG PP3/BP4 evidence strength within MANE Select transcripts** | | | | | | | | |
| --- | --- | --- | --- | --- | --- | --- | --- | --- |
| **ACMG/AMP evidence code** | **Computational tool** | | | | | | | |
|  | **BayesDel** | | **MutPred2** | | **REVEL** | | **VEST4** | |
|  | Median (range) | Mean  (sd) | Median (range) | Mean  (sd) | Median (range) | Mean  (sd) | Median (range) | Mean  (sd) |
| PP3_Strong | 1  (0-4) | 1.1  (1) | 1  (0-7) | 1  (1) | 1  (0-4) | 0.8  (0.9) | 1  (0-5) | 1.1  (1.1) |
| PP3_Moderate | 5  (1-10) | 4.8  (2.1) | 5  (0-14) | 5.6  (2.4) | 4  (0-11) | 4.5  (2.3) | 6  (0-16) | 5.9  (2.3) |
| PP3_Supporting | 6  (1-14) | 5.8  (2.4) | 6  (1-14) | 6.2  (2.6) | 5  (1-13) | 5.1  (2.2) | 6  (0-16) | 6.6  (2.9) |
| Indeterminate | 29  (15-57) | 29  (6.6) | 33  (15-67) | 33.9  (7.9) | 30  (16-61) | 30.2  (7.4) | 24.5  (10-60) | 25.7  (7) |
| BP4_Supporting | 32  (18-74) | 34.1  (9.2) | 35  (19-82) | 36.8  (9.6) | 22  (11-55) | 22.5  (6.8) | 18  (8-51) | 19.8  (6.7) |
| BP4_Moderate | 73  (51-187) | 77.6  (23.4) | 64  (45-172) | 69.4  (21.4) | 60  (41-144) | 64.1  (18.3) | 78  (54-178) | 82.4  (21) |
| BP4_Strong | - | - | 0  (0-3) | 0.3  (0.6) | 3  (0-13) | 3.4  (2.2) | - | - |
| BP4_Very Strong | - | - | - | - | 0  (0-2) | 0.1  (0.4) | - | - |
| No score | 0  (0-4) | 0.7  (0.8) | 0  (0-0) | 0  (0) | 21  (8-51) | 22.5  (7.4) | 11  (3-31) | 11.8  (4.7) |

“-” indicates that the given tool is not able to provide BP4 evidence of this strength.

| **Table S4. Number of rare (≤1% AF) missense variants per proband in disease-associated genes by ACMG PP3/BP4 evidence strength using VEP “most severe consequence”** | | | | | | | | |
| --- | --- | --- | --- | --- | --- | --- | --- | --- |
| **ACMG/AMP evidence code** | **Computational tool** | | | | | | | |
|  | **BayesDel** | | **MutPred2** | | **REVEL** | | **VEST4** | |
|  | Median (range) | Mean  (sd) | Median (range) | Mean  (sd) | Median (range) | Mean  (sd) | Median (range) | Mean  (sd) |
| PP3_Strong | 1  (0-4) | 1.1  (1) | 1  (0-4) | 0.8  (0.9) | 1  (0-4) | 0.8  (0.9) | 1  (0-4) | 0.9  (0.9) |
| PP3_Moderate | 4  (0-10) | 4.2  (1.9) | 4  (0-13) | 4.1  (2) | 3  (0-10) | 3.7  (2) | 5  (0-11) | 5.1  (2) |
| PP3_Supporting | 5  (0-13) | 4.8  (2.2) | 4  (0-11) | 4  (2.1) | 4  (0-10) | 4.2  (2) | 5  (0-14) | 5.5  (2.4) |
| Indeterminate | 19  (8-36) | 19.7  (5.1) | 20  (9-50) | 21.1  (5.5) | 21  (9-47) | 22.1  (5.8) | 19  (5-33) | 19.2  (4.6) |
| BP4_Supporting | 19  (10-54) | 19.5  (5.5) | 20  (10-43) | 20.4  (5.5) | 13  (5-29) | 13.6  (4.2) | 12  (3-42) | 12.2  (4.4) |
| BP4_Moderate | 36  (20-101) | 37.6  (11) | 39  (21-116) | 40.2  (11.4) | 35  (18-107) | 37.2  (10.4) | 36  (19-103) | 37.3  (11.2) |
| BP4_Strong | - | - | 0  (0-2) | 0.1  (0.3) | 2  (0-12) | 2.2  (1.7) | - | - |
| BP4_Very Strong | - | - | - | - | 0  (0-2) | 0.1  (0.3) | - | - |
| No score | 4  (0-11) | 3.9  (2) | 0  (0-0) | 0  (0) | 6  (1-17) | 6.8  (2.7) | 10  (3-29) | 10.5  (3.8) |

“-” indicates that the given tool is not able to provide BP4 evidence of this strength.

| **Table S5. Number of rare (≤1% AF) missense variants per proband genome-wide by ACMG PP3/BP4 evidence strength within MANE Select transcripts** | | | | | | | | |
| --- | --- | --- | --- | --- | --- | --- | --- | --- |
| **ACMG/AMP evidence code** | **Computational tool** | | | | | | | |
|  | **BayesDel** | | **MutPred2** | | **REVEL** | | **VEST4** | |
|  | Median (range) | Mean  (sd) | Median (range) | Mean  (sd) | Median (range) | Mean  (sd) | Median (range) | Mean  (sd) |
| PP3_Strong | 2  (0-6) | 2.1  (1.3) | 3  (0-8) | 2.8  (1.6) | 1  (0-6) | 1.5  (1.3) | 3  (0-9) | 2.7  (1.6) |
| PP3_Moderate | 11  (3-22) | 10.9  (3.3) | 13  (5-25) | 13.1  (3.5) | 8  (2-18) | 8.5  (3) | 16  (5-29) | 16.4  (4.4) |
| PP3_Supporting | 14  (6-27) | 14  (4) | 13.5  (3-24) | 13.8  (3.7) | 10  (2-19) | 10  (3.1) | 17  (7-30) | 17.5  (4.1) |
| Indeterminate | 60  (39-112) | 60.8  (10.9) | 71  (47-137) | 72.8  (12.9) | 58  (39-114) | 59.6  (10.9) | 70  (44-127) | 70.6  (12.4) |
| BP4_Supporting | 65  (41-174) | 67.4  (14.7) | 71  (47-191) | 74.4  (16.3) | 44.5  (24-104) | 45.5  (9.4) | 46  (23-127) | 48.2  (10.8) |
| BP4_Moderate | 167  (115-507) | 176.7  (42.1) | 150  (104-476) | 159.3  (39.2) | 151  (103-470) | 159.3  (37.4) | 154  (103-488) | 164.6  (40.7) |
| BP4_Strong | - | - | 1  (0-5) | 0.9  (1) | 12  (3-35) | 12.6  (4.6) | - | - |
| BP4_Very Strong | - | - | - | - | 0  (0-4) | 0.7  (0.8) | - | - |
| No score | 5  (0-15) | 5.3  (2.6) | 0  (0-0) | 0  (0) | 38  (22-118) | 39.6  (10.6) | 16  (6-57) | 17.3  (6) |

“-” indicates that the given tool is not able to provide BP4 evidence of this strength.
